# Mental healthcare and service user impact of the COVID-19 pandemic: results of a UK survey of staff working with people with intellectual disability and developmental disorders

**DOI:** 10.1101/2020.09.01.20178848

**Authors:** Rory Sheehan, Christian Dalton-Locke, Afia Ali, Vasiliki Totsika, Norha Vera San Juan, Angela Hassiotis

**Affiliations:** Division of Psychiatry, University College London, London, UK; NIHR Mental Health Policy Research Unit, Division of Psychiatry, University College London, London, UK; NIHR Mental Health Policy Research Unit, Instituted of Psychiatry, Psychology & Neuroscience, King’s College London, London, UK

**Keywords:** COVID-19, Coronavirus, intellectual disabilities, developmental disorders, mental health services

## Abstract

**Background:** Very little is known about the impact of previous epidemics on the care of people with intellectual and developmental disabilities, particularly in terms of mental health services. The COVID-19 pandemic has the potential to exacerbate existing health inequalities as well as expose gaps in service provision for this vulnerable population group.

**Methods:** We investigated the responses of 648 staff working in mental healthcare with people with intellectual disabilities and/or developmental disabilities. Participants contributed to a UK-wide online survey undertaken by the National Institute for Health Research Mental Health Policy Research Unit between 22^nd^ April and 12^th^ May 2020. Recruitment was via professional networks, social media and third sector organisations. Quantitative data describing staff experience over three domains (challenges at work, service user and carer problems, sources of help at work) were summarised and differences between groups explored using Chi square tests. Content analysis was used to organise qualitative data focusing on service changes in response to the pandemic.

**Results:** The majority of survey respondents worked in the NHS and in community mental health services. One third had managerial responsibility. Major concerns expressed by mental healthcare staff were: difficulties for service users due to lack of access to usual support networks and health and social care services during the pandemic; and difficulties maintaining adequate levels of support secondary to increased service user need. Staff reported having to quickly adopt new digital ways of working was challenging; nevertheless, free text responses identified remote working as the innovation that staff would most like to retain after the pandemic subsides.

**Conclusions:** Understanding the experiences of staff working across different settings in mental healthcare for people with intellectual and developmental disabilities during the COVID-19 pandemic is essential in guiding contingency planning and fostering service developments to ensure the health of this vulnerable group is protected in any future disease outbreaks.

## INTRODUCTION

The World Health Organisation first announced the COVID-19 outbreak as a Public Health Emergency of International Concern on 30^th^ January 2020 and subsequently upgraded it to a pandemic on 11^th^ March. Several measures were taken to combat the spread of the infection including, social distancing, shielding of vulnerable individuals, restrictions on many sectors of the economy, and advice to stay at and/or work from home, where possible. This unprecedented national ‘lockdown’ represented the most drastic change to life in several decades and has had far-reaching implications. The negative short and longer-term psychological effects of quarantine on population mental health have been shown in a recent systematic review of studies of previous infectious disease outbreaks, and include post-traumatic stress disorder, confusion, and anger (Brooks et al. 2020).

For people with intellectual and other developmental disabilities such as autism, (approximately 3-5 in 100 of the population (Srivastava and Schwartz 2014), the pandemic presents very specific challenges. In addition to being at increased risk of mental health problems (Hughes-McCormack et al. 2018), people with intellectual disability are a group with significant health comorbidity, including frailty, obesity, diabetes, and respiratory disease (Cooper et al. 2015), which makes them immediately more vulnerable worse outcomes of infection with COVID-19 (Turk et al. 2020). Furthermore, people with intellectual and developmental disabilities experience severe health inequalities including early death from preventable causes (LeDeR, 2020; Hirvikoski et al. 2016).

A substantial proportion of adults with intellectual and other developmental disabilities live in congregate settings or supported housing, sharing with others and being dependant on staff for aspects of their day-to-day care. Such services, particularly care homes, are vulnerable to the rapid spread of infection and worse outcomes (Landes et al. 2020). Other concerns of particular relevance to this population group are the destabilising effect of service disruptions and breaks in routine, inability to follow social distancing measures or to wear personal protective equipment, and difficulty in coping with isolation and lack of contact with family, friends, and known trusted staff (Courtenay and Perera 2020).

The pandemic has forced a reduction in face-to-face routine appointments and a simultaneous increase in urgent and emergency presentations (Royal College of Psychiatrists 2020). Staff working in mental health and intellectual disability services have rapidly adopted new ways of working in an attempt to balance continuity of care delivery with reduction in transmission of infection. These changes are, in the main, untested. Working from home, for example, may pose challenges, especially where staff have caring responsibilities or have had little preparation and guidance. Equally untested is the engagement of services users with intellectual and other developmental disabilities in remote care.

In this paper we report a secondary analysis of the experiences of staff working within a variety of mental health services for people with intellectual and other developmental disabilities. The findings reveal gaps in mental health and social care for people with intellectual and developmental disabilities and should inform future directions for service delivery and research to improve support during subsequent waves of the current pandemic or future public health emergencies.

## METHODS

The original study (Johnson et al. 2020) was approved by the King’s College London Research Ethics Committee (MRA-19/20-18372). A survey was developed by staff, academics and people with relevant lived experience working as part of the National Institute for Health Research (NIHR) Mental Health Policy Research Unit. An expert in intellectual disabilities (AH) provided input into questions relating to people with intellectual and other developmental disabilities. The survey aimed to include professionals from NHS mental health and social care services, along with those working in the private and third sectors. Following internal piloting, the survey was made publicly available on the Opinio platform and was disseminated via multiple channels including the Royal College of Psychiatrists and the Royal College of Nurses, trade unions, social media, academic interest groups, housing providers, and social care and voluntary sector organisations. Additional efforts were made to increase the inclusion of Black, Asian and Minority Ethnic Group staff in the sample. The survey was open during the mid stage of UK lockdown restrictions (22 April to 12 May 2020).

The survey comprised three main sections covering, (1) challenges at work during the COVID-19 pandemic, (2) staff perspectives of problems faced by mental health service users and family carers and, (3) sources of help at work in managing the impact of the pandemic. Respondents who indicated that they worked with people with intellectual and other developmental disabilities were filtered to an additional set of questions specifically relevant to this group. Most of the survey items were in the form of short statements which respondents rated on a five-point Likert scale between “not relevant/not important” and “extremely relevant/extremely important”. The questionnaire also included a number of open-ended questions seeking information about service innovations and adaptations, their perceived success, concerns about the future, and which new ways of working should be maintained after the pandemic.

### Analysis

Descriptive statistics were used to summarise participant characteristics and responses across each of the three main sections of the survey. We report the percentage of the sample who rated each survey item “very relevant/very important” or “extremely relevant/extremely important”. In order to explore potential differences in experiences between different groups of respondents, we categorised the responses to the challenges at work questions by sector (staff working in NHS or non-NHS sectors), managerial responsibility (yes or no), and setting (staff working in community or in-patient settings). We created a binary variable for each survey item formed of “not relevant” and “any relevant” responses and used Chi square tests to examine group differences. We applied a stringent alpha of 0.001 to avoid type 1 error due to multiple testing. We carried out a sensitivity analysis including responses only from those who worked with people with intellectual and other developmental disability as respondents could indicate that they worked with multiple service user groups. We believe that as people with intellectual and other developmental disabilities often use generic mental health services, staff have legitimate reasons to score items based on their experience with this population. All analyses were conducted using Stata v15 (StataCorp 2017).

As we gradually move towards the relaxation of lockdown measures and attention turns to future planning, we have focused our qualitative analysis on service innovations that may be of relevance moving forward. Content analysis was used to analyse free text responses to the question “Has any innovation or change been made in mental healthcare that you would like to remain in place after the pandemic subsides?” Researchers initially familiarised themselves with the data and then developed codes to report the qualitative data in more succinct representative categories (Hsieh and Shannon 2005). To enhance credibility, two researchers (VT and AA) worked in parallel to explore the unedited participant responses and identify emerging themes and sub-themes, using Microsoft Excel to organise the data. The final coding frame was developed through ongoing discussion between them and with the whole team, and applied independently by the two researchers on 25% of randomly selected participant responses. Inter-rater reliability was good (78%), therefore the remaining data were indexed by one researcher (VT). Prevalence of themes in the data was separated by the two most sizeable sub-groups of survey respondents and in keeping with the quantitative subgroup analysis (NHS vs non-NHS staff and managers vs not managers). Themes are illustrated with brief direct quotations.

## RESULTS

3,172 staff started the survey. Of these, 902 (28.4%) indicated they worked with people with intellectual and/or developmental disabilities. The results that follow include those who answered at least one question from each of the three core domains that were open to all respondents (*n=*648).

### Participant characteristics

The large majority of respondents were female (*n=*401, 78.9%), aged 25-64 years (*n=*471, 92.7%) and of white ethnicity (*n=*421, 82.2%). Respondents had been working in mental health services for a mean of 14.5 years (standard deviation 10.5 years). The majority were based in England (*n=*526, 81.3%) with smaller proportions in Scotland (*n=*69, 10.6%), Wales (*n=*39, 6%), and Northern Ireland (*n=*6, 0.9%).

About a third of respondents reported caring for children (*n=*158, 31%) and a quarter for elderly relatives (*n=*135, 26.7%). A third were working from home only (*n=*172, 33.9%), just over a third were at the workplace (*n=*178, 35.1%) and the remainder (*n=*147, 28.9%) worked from both home and at the workplace (a small number were not working due to sickness or self-isolating when they completed the survey). The majority worked in the NHS (*n=*539, 83.1%), with a minority working in social care, and the voluntary or private sectors. Just over half worked in community settings (*n=*373, 57.6%). 182 were nurses (28.2%), 104 were psychologists (16.1%), 55 were psychiatrists (8.5%), 40 were social workers (6.2%), and 109 were support workers (16.9%). One third of respondents indicated they had management responsibilities (*n=*230, 35.5%). Full demographic details, including missing data for each item, are shown in data table 1.

**Table 1.** Demographic and employment characteristics of survey respondents (*n*=648)

|  |  | *n* | Valid % |
| --- | --- | --- | --- |
| **Demographic variables** |  |  |  |
| Sex | Male | 96 | 18.9 |
|  | Female | 401 | 78.9 |
|  | Other | 2 | 0.4 |
|  | Prefer not to say | 9 | 1.8 |
|  | Missing | 140 | - |
| Age group | <25 | 21 | 4.1 |
|  | 25-34 | 127 | 25.0 |
|  | 35-44 | 90 | 17.7 |
|  | 45-54 | 155 | 30.5 |
|  | 55-64 | 99 | 19.5 |
|  | ≥65 | 3 | 0.6 |
|  | Prefer not to say | 13 | 2.6 |
|  | Missing | 140 | - |
| Ethnic group | White | 421 | 86.6 |
|  | Asian | 26 | 5.3 |
|  | Black | 12 | 2.5 |
|  | Multiple groups / mixed | 20 | 4.1 |
|  | Other | 4 | 0.8 |
|  | Prefer not to say | 3 | 0.6 |
|  | Missing | 162 | - |
| Caring for children <18 years | Yes | 158 | 31.1 |
|  | No | 350 | 68.9 |
|  | Missing | 140 | - |
| Caring for elderly or disabled relative | Yes | 135 | 26.8 |
|  | No | 369 | 73.2 |
|  | Missing | 144 | - |
| **Employment variables** |  |  |  |
| Work sector* | NHS | 539 | - |
|  | Social care | 52 | - |
|  | Voluntary | 43 | - |
|  | Community or user-led | 11 | - |
|  | Private | 28 | - |
| Work setting* | Hospital in-patient | 161 | - |
|  | Crisis house in community | 7 | - |
|  | Residential / supported accommodation | 40 | - |
|  | Crisis team, liaison team, AMHP team | 108 | - |
|  | Community team | 373 | - |
|  | Day/drop-in/recovery college | 51 | - |
|  | Other | 99 | - |
| Professional role | Psychologist | 104 | 16.1 |
|  | Nurse | 182 | 28.2 |
|  | Occupational therapist | 42 | 6.5 |
|  | Other qualified therapist | 75 | 11.6 |
|  | Peer support worker | 21 | 3.3 |
|  | Psychiatrist | 55 | 8.5 |
|  | Social worker | 40 | 6.2 |
|  | Manager, no mental health professional qualification | 18 | 2.8 |
|  | Other worker with direct contact with people with mental health problems | 109 | 16.9 |
|  | Missing | 2 | - |
| Managerial responsibility | Yes | 230 | 35.5 |
|  | No | 418 | 64.5 |
|  | Missing | 0 | - |
| Which country do you work in | England | 526 | 81.3 |
|  | Northern Ireland | 6 | 0.9 |
|  | Scotland | 69 | 10.7 |
|  | Wales | 39 | 6.0 |
|  | Other | 7 | 1.1 |
|  | Missing | 1 | - |
| What locality do you work in | City/town >100,000 population | 437 | 67.8 |
|  | Town < 100,000 population | 142 | 22.0 |
|  | Rural | 66 | 10.2 |
|  | Missing | 3 | - |

\*Percentages not given as respondents were able to select more than one response

### Quantitative results

The results reported below are based on respondents who rated items as “very relevant/very important” or “extremely relevant/extremely important”. The most highly rated items are listed in the narrative with further details available in data tables 2, 3, and 4.

**Table 2.** Ranking of current challenges at work items

| Rank | Item | Valid responses | %very or extremely relevant |
| --- | --- | --- | --- |
| 1 | Having to adapt too quickly to new ways of working | 644 | 53.3 |
| 2 | Increased need following the withdrawal of educational and support services in the community | 395 | 52.2 |
| 3 | Difficulty maintaining adequate levels of support for those with significant and complex needs | 399 | 50.1 |
| 4 | Difficulty for service users in comprehending the current crisis and the resulting requirements | 398 | 49.7 |
| 5 | Difficulty engaging people with ID and/or autism with remote appointments | 395 | 45.3 |
| 6 | Difficulty maintaining adequate support for families looking after a child/young person/adult with ID and/or autism who displays challenging behaviour | 394 | 42.9 |
| 7 | Having to respond to additional mental health needs that appear to result from COVID19 | 647 | 40.8 |
| 8 | Service users no longer getting an acceptable service due to service reconfiguration due to COVID19 | 645 | 38.6 |
| 9 | The risk I or my colleagues could be infected with COVID19 at work | 641 | 38.4 |
| 10 | Having to learn to use new technologies too quickly and/or without sufficient training and support | 644 | 37.9 |
| 11 | Pressures resulting from the need to support colleagues through the stresses associated with the pandemic | 643 | 35.9 |
| 12 | The risk that COVID19 will spread between service users I'm working with | 643 | 35.0 |
| 13 | Concern that physical healthcare received by service users I work with may not be adequate | 642 | 34.7 |
| 14 | Being expected to use new technologies without reliable access to necessary tools and equipment | 644 | 34.3 |
| 15 | Concerns about discrimination in access to physical health care for COVID19 | 394 | 45.3 |
| 16 | Lack of high quality and relevant information for people with ID and/or autism about the COVID pandemic and the requirements that result | 398 | 33.4 |
| =17 | The risk family or friends may be infected with COVID19 through me | 647 | 32.8 |
| =17 | Problems resulting from lack of access to testing | 644 | 32.8 |
| 19 | Difficulty putting infection control measures into practice in the setting I work in | 644 | 29.3 |
| 20 | Greater workload than usual | 648 | 27.9 |
| 21 | Lack of protective clothing (PPE) and equipment needed for infection control | 644 | 25.8 |
| 22 | Safeguarding and other risk management processes cannot be adequately mobilised due to limited social care, legal or police response | 646 | 23.7 |
| 23 | Increased difficulty managing work-life balance, for example because of loss of childcare | 645 | 23.4 |
| 24 | Working longer hours than usual | 647 | 21.5 |
| 25 | Staff shortages (more than is usual in this setting) | 645 | 20.8 |
| 26 | Working in a different clinical setting or with different clinical problems from usual | 646 | 20.0 |
| 27 | Feeling less able to do my job than usual because my own well-being has suffered through the stresses of the pandemic | 646 | 17.5 |
| 28 | Feeling under pressure from managers or colleagues to be less cautious about infection control than I would like | 647 | 15.0 |
| 29 | Not enough of the team I'm working with are permanently employed in this setting (lots of bank/locum and redeployed staff) | 648 | 10.3 |
| 30 | Pressure to accept redeployment to a setting where I don't feel happy to work | 646 | 9.4 |
| 31 | Problems commuting safely to work and back | 648 | 8.6 |

**Table 3.** Ranking of service user and carer problem items

| Rank | Item | Valid responses | %very or extremely important |
| --- | --- | --- | --- |
| 1 | Lack of access to usual support networks of family and friends | 645 | 73.6 |
| 2 | Lack of usual work and activities | 644 | 69.6 |
| 3 | Loneliness due to or made worse by social distancing, self-isolating, or shielding | 648 | 69.4 |
| 4 | Lack of access to usual support from other services (primary care, social care, voluntary sector) | 647 | 62.4 |
| 5 | Worries about getting COVID-19 infection | 648 | 61.1 |
| 6 | Worries about family getting COVID-19 infection | 647 | 59.2 |
| 7 | Increased difficulties for families / carers | 643 | 56.3 |
| 8 | Lack of access to usual support from NHS mental health services | 645 | 52.4 |
| 9 | Relapse and deterioration in mental health triggered by COVID-19 stresses | 648 | 47.2 |
| 10 | High personal risk of severe consequences of COVID-19 infection (e.g. due to comorbidities) | 643 | 46.5 |
| 11 | Difficulty understanding or following current government requirements on social distancing, self-isolation, shielding | 644 | 43.3 |
| 12 | Increase in reliance on family / family tensions | 644 | 42.7 |
| 13 | Difficulty engaging with remote appointments by phone or via digital platforms | 642 | 38.5 |
| 14 | Increased risk from abusive domestic relationships | 645 | 38.0 |
| 15 | Diminished access to physical health care for problems other than COVID-19 | 647 | 35.6 |
| 16 | Having to stay at home in poor circumstances or not having a home to go to | 646 | 33.8 |
| 17 | Difficulty getting food, money, or other basic resources | 642 | 33.0 |
| 18 | Risk of increased drug and alcohol use or gambling | 644 | 28.1 |
| 19 | Effects of COVID-19-related trauma | 644 | 27.5 |
| 20 | Lack of access to or of equitable provision of physical healthcare for COVID-19 | 647 | 22.7 |
| 21 | Loss of liberty and rights due to changes in implementation of mental health legislation | 645 | 19.4 |
| 22 | Lack of access to medication and to processes for administering and monitoring it | 646 | 18.4 |
| 23 | Problems with police or other authorities because of lack of understanding / ability to stick to current government requirements | 644 | 14. |

**Table 4.** Ranking of sources of support items

| Rank | Item | Valid responses | %very or extremely relevant |
| --- | --- | --- | --- |
| 1 | Guidance from my employer on managing clinical and safety needs due to COVID-19 | 648 | 65.9 |
| 2 | Support and information from colleagues | 645 | 65.7 |
| 3 | Resilience and resourcefulness in adversity among service users and carers | 648 | 65.3 |
| 4 | Support and advice from my manager(s) | 644 | 64.1 |
| 5 | Adoption of new digital ways of working | 639 | 60.7 |
| 6 | Guidance disseminated by the NHS or professional bodies | 641 | 58.5 |
| 7 | Being aware of public support for key workers | 645 | 45.1 |
| 8 | Staff well-being initiatives set up during COVID-19 in my workplace | 644 | 41.3 |
| 9 | The support offered by local volunteers and mutual aid groups | 643 | 37.8 |
| 10 | National initiatives to support service users and carers, such as helplines or online peer support | 639 | 37.3 |
| 11 | Support and new initiatives from local voluntary sector organisations | 642 | 36. |
| 12 | New initiatives in NHS mental health services | 641 | 35.4 |
| 13 | National initiatives to support staff well-being | 645 | 32.7 |
| 14 | Information from the media or social media | 647 | 22.7 |

**Table 5.**
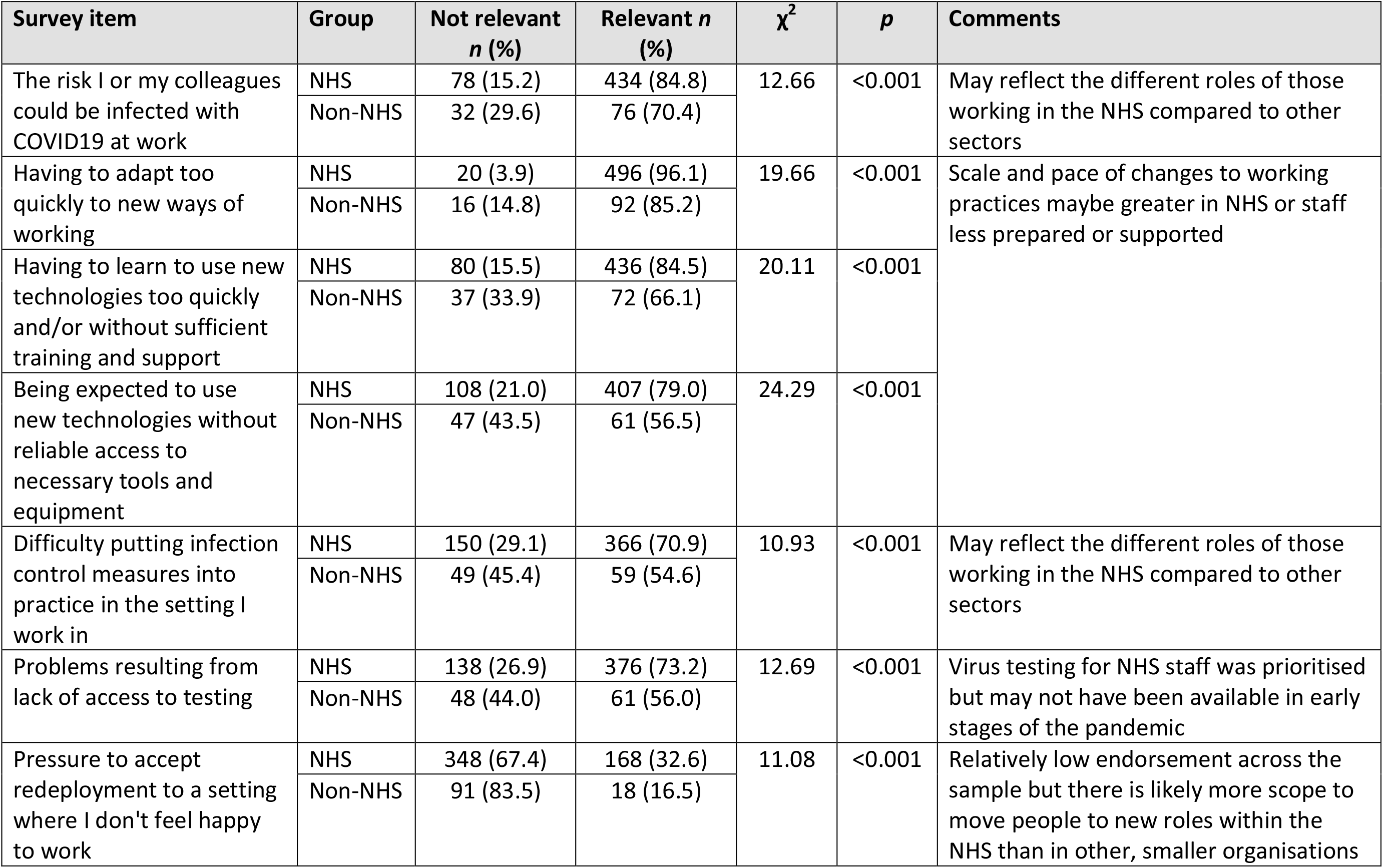
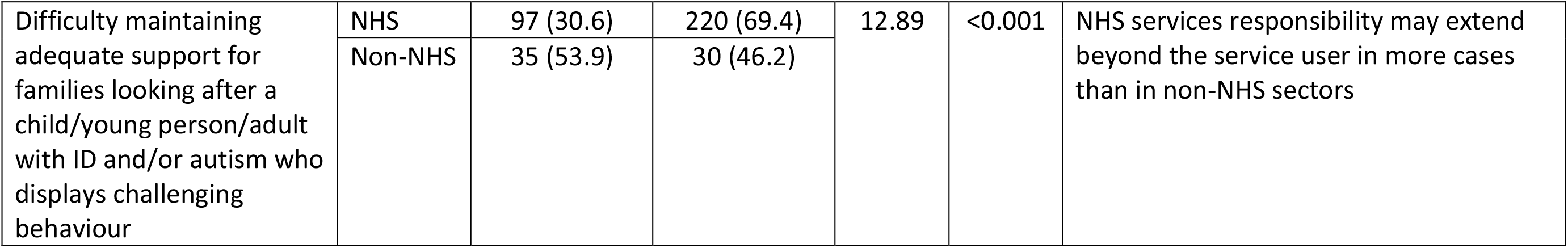
NHS and non-NHS staff – significant differences in staff reporting items as relevant or not relevant

**Table 6.**
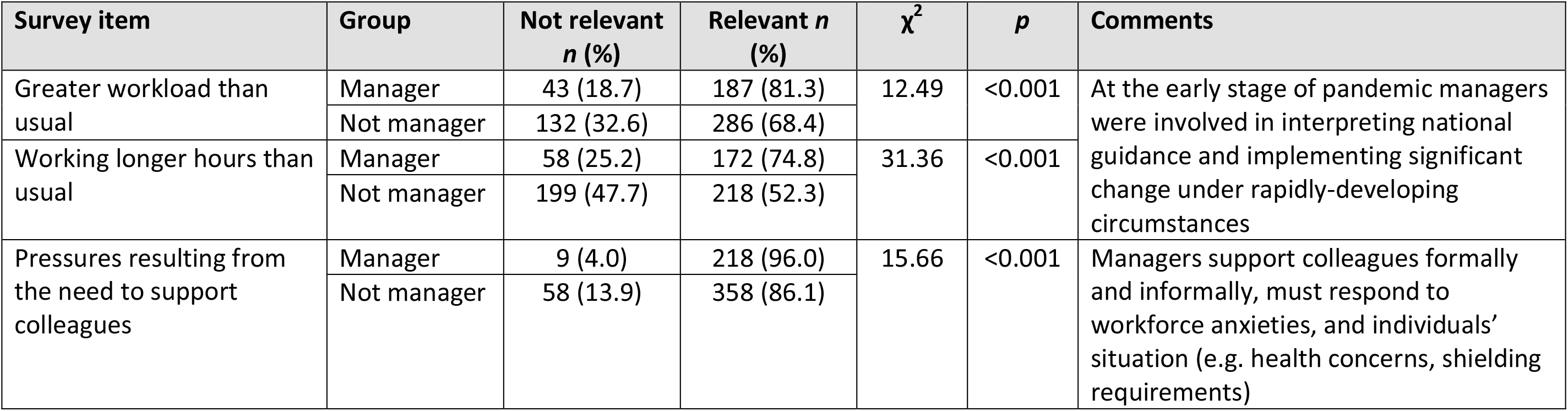
Managers and non-managers – significant differences in staff reporting items as relevant or not relevant

| Survey item | Group | Not relevant<br>*n* (%) | Relevant *n*<br>(%) | $\chi^2$ | *p* | Comments |
| --- | --- | --- | --- | --- | --- | --- |
| Greater workload than usual | Manager | 43 (18.7) | 187 (81.3) | 12.49 | <0.001 | At the early stage of pandemic managers were involved in interpreting national guidance and implementing significant change under rapidly-developing circumstances |
|  | Not manager | 132 (32.6) | 286 (68.4) |  |  |  |
| Working longer hours than usual | Manager | 58 (25.2) | 172 (74.8) | 31.36 | <0.001 |  |
|  | Not manager | 199 (47.7) | 218 (52.3) |  |  |  |
| Pressures resulting from the need to support colleagues | Manager | 9 (4.0) | 218 (96.0) | 15.66 | <0.001 | Managers support colleagues formally and informally, must respond to workforce anxieties, and individuals' situation (e.g. health concerns, shielding requirements) |
|  | Not manager | 58 (13.9) | 358 (86.1) |  |  |  |

**Table 7.**
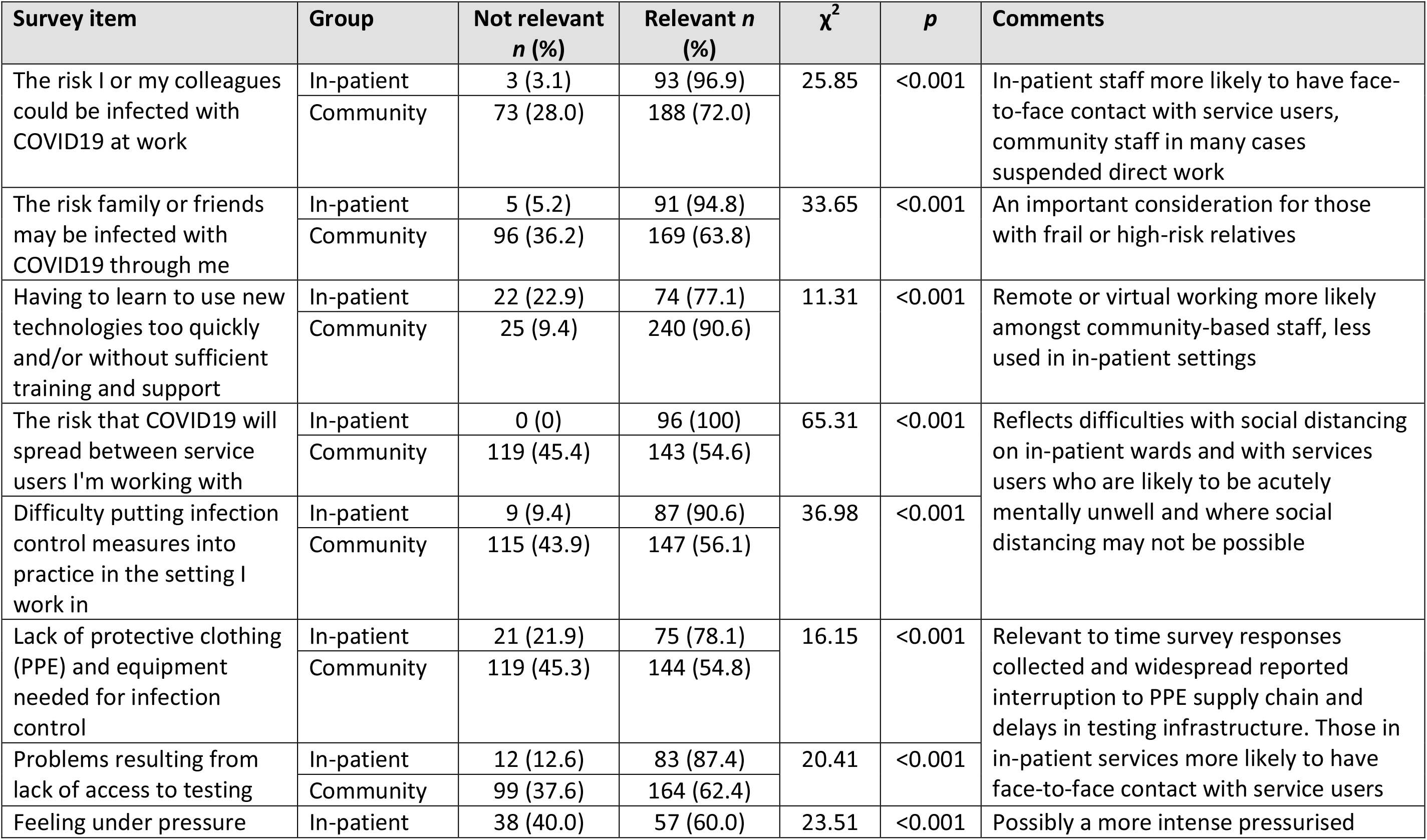
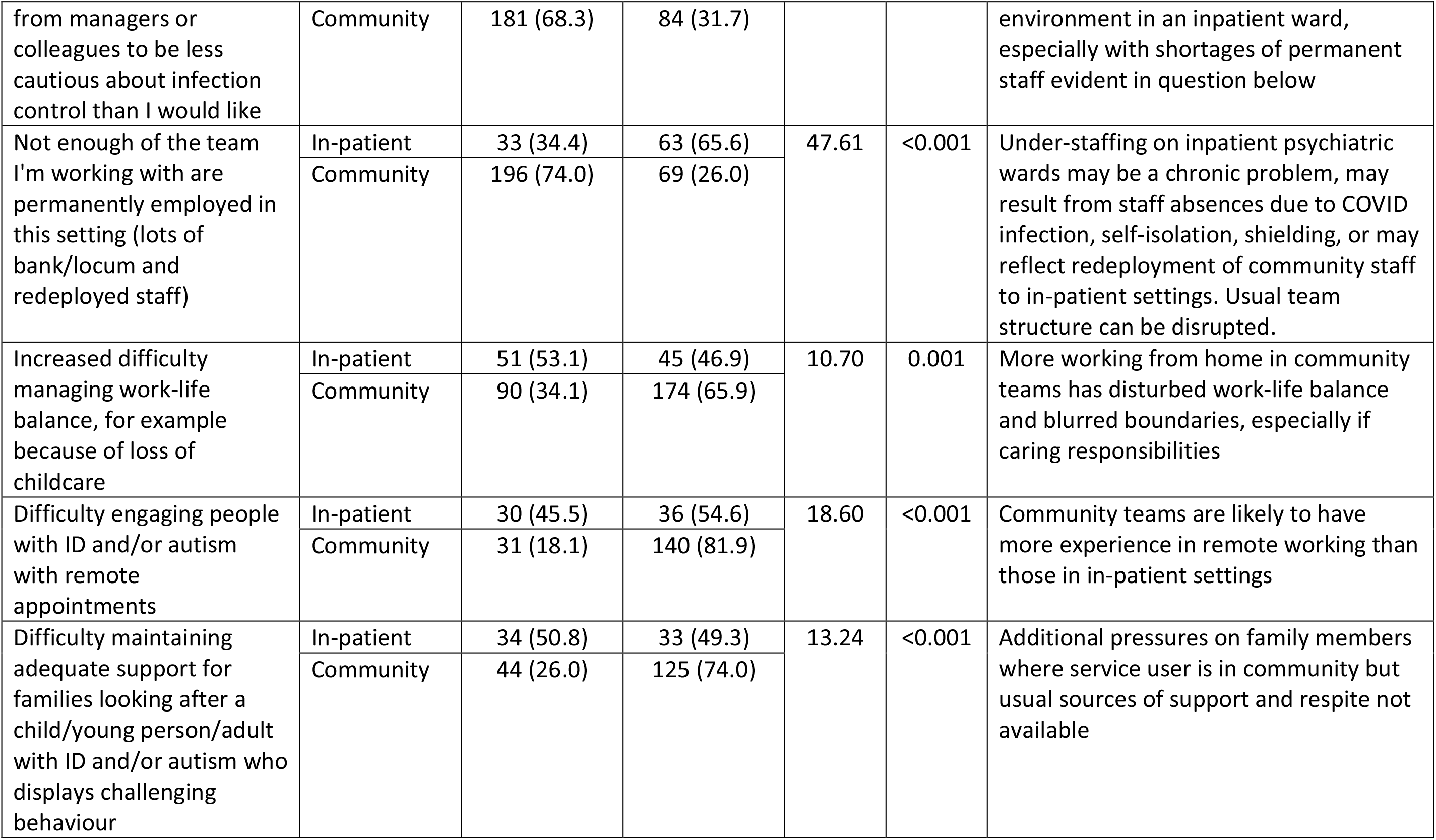
In-patient and community staff – significant differences in staff reporting items as relevant or not relevant

#### Challenges at work

Staff rated as most relevant the following challenges: having to adapt too quickly to new ways of working (rated as very or extremely relevant by 53.3% respondents), increased service user need following withdrawal of educational and support services in the community (52.2% respondents), and difficulty maintaining adequate levels of support for those with significant and complex needs (50.1% respondents). These concerns were closely followed by concerns that service users had difficulty in comprehending the crisis and the resulting requirements (49.7%) and in engaging people with intellectual and other developmental disabilities with remote appointments (45.3%).

#### Staff perception of service users’ and carers’ problems

Staff perception of the concerns of service users and carers centred mainly on the loss of usual supports and routine secondary to the lockdown restrictions. Lack of access to support networks of family and friends (rated as very or extremely relevant by 73.6% respondents), pause of work or activities (69.6% respondents), and of usual support from other services (e.g. primary care, social care, voluntary sector) (62.4%) were all seen as relevant fuelling fears that loneliness would be made worse by social distancing, self-isolation, or shielding (69.4% respondents).

#### Sources of help at work

The most important sources of help at work were as follows: guidance from employer (65.9% respondents); support and information from colleagues (65.7% respondents); and resilience and resourcefulness among service users and carers (65.3%). Being aware of public support for keyworkers was rated as important by 45.1% respondents.

### Qualitative results

Three hundred and sixty-four respondents answered the free text question that we analysed. Three main themes were identified in the responses: remote operation, flexibility, and organisational improvement.

**Remote operation** included remote staff working and remote service provision as two distinct sub-themes. These sub-themes referred to the ability to work from home and using remote technology to communicate, provide therapy, and maintain contact with clients. Remote operation was considered positive because it is more immediate, faster, can improve work-life balance, and is better for the environment. Respondents proposed that remote services for those in need can reduce waiting times and the rate of non-attendance.

> “*As a carer, more home working has given me a better work-life balance…the reduced stress means I am able to focus more on patient needs”*
>
> *“Telephone and online counselling for some clients has been beneficial as they struggle to access the building…my DNA [did not attend] rate has decreased as a result”*

A related theme to that of remote operation was that of **flexibility** in the way professionals work, and in the services offered to patients. The latter can choose whether s/he prefers an ‘in person’ or remote consultation. It involves managers monitoring staff productivity by task and outcome, rather than hours spent in the office. Flexibility was thought to improve productivity, resulting in more efficient working.

> “*Finally some flexibility!”*
>
> *“[I’ve noticed] a change in attitude towards agile working - task oriented rather than a focus on time spent working”*

**Organisational improvement** referred to specific descriptions of organisational changes that participants identified. Within organisational improvement, sub-themes were identified which referred to the development of a new service, initiatives focused on staff wellbeing, the integration of services, and improved communication between staff and leadership style from managers (see table 8).

> “*More thinking outside the box…a more ‘hands-on’ approach”*
>
> *“Colleagues being very caring and looking after one another…the team has really pulled together for clients’ and staff wellbeing”*
>
> *“E-mails from senior managers and directors have taken a much more humane tone, and seem far more genuine than the usual bland, generic, corporate speak”*

**Table 8.** Innovations and changes made during the COVID-19 pandemic that survey respondents would like to retain – content analysis

|  | Sector |  | Managerial responsibility |  |
| --- | --- | --- | --- | --- |
|  | NHS | Non-NHS | Managers | Non-managers |
| Valid answers | 314 | 50 | 150 | 219 |
| **Remote operation** |  |  |  |  |
| Remote working for staff | 48% | 36% | 48% | 44% |
| Remote services for service users | 51% | 42% | 51% | 40% |
| **Flexibility** | 7% | 8% | 7% | 10% |
| **Organisational improvement** | 17% | 18% | 15% | 18% |
| New services | 7% | 4% | 6% | 8% |
| Staff well-being | 6% | 4% | 5% | 7% |
| Service integration | 1% | 2% | 1% | 0.5% |
| Better leadership | 1% | 2% | 1% | 2% |

NB - Figures are the percentage of respondents in each group who mentioned each theme or sub-theme

### Group differences in quantitative and qualitative results patterns

#### NHS vs non-NHS staff

Those working in NHS settings were more concerned about being infected with COVID-19 whilst at work than those working in non-NHS settings (percentage rating this item as relevant 84.8% vs 70.4%, p<0.001). Staff working in NHS settings expressed more problems with implementing precautions designed to minimise the transmission of infection, including: lack of personal protective equipment (65.7% vs 52.3%, p=0.009); difficulty putting infection control measures into place (70.9% vs. 54.6%, p<0.001); and problems resulting from lack of access to testing (73.2% vs 56.0%, p<0.001).

Having to adapt too quickly to new ways of working (96.1% vs 85.2%, p<0.001), having to use new technologies without adequate training or support (84.5% vs 66.1%, p<0.001), and not having necessary tools or equipment to facilitate remote working (79.0% vs 56.5%, p<0.001) were all significantly more relevant to NHS staff compared to those working in other sectors.

Despite these difficulties in remote and digital working, the qualitative results demonstrated that remote operation was the service innovation selected most frequently by those working in NHS and non-NHS settings: remote staff working was mentioned by 48% of NHS staff and 36% of non-NHS staff, while remote service provision was identified by 52% of NHS staff and 36% of non-NHS staff. Flexibility and organisational improvements were service innovations mentioned less frequently by participants in either group (table 8).

#### Management vs non-management staff

Survey respondents who had managerial responsibility were more likely to endorse having a greater workload than usual (81.3% vs 68.4%, p<0.001) and to be working longer hours (74.8% vs 52.3%, p<0.001) than those without a management role. They also felt more pressure arising from the need to support colleagues through the pandemic (96.0% vs 86.1%, p<0.001).

About half of those with managerial responsibility mentioned remote working (48%) and remote service provision (51%) as the service innovation they were most likely to want to retain after the pandemic, slightly higher than those without managerial responsibility (40% and 44%, respectively). Flexibility and organisational improvements were service innovations mentioned less frequently by participants in either group (table 8).

### Community vs in-patient staff

Staff working in-patient settings were more concerned about infection risk to themselves, to their family/friends through them, and the spread of infection between service users compared with those working in community settings (corresponding percentages 96.9% vs 72.0%, 94.8% vs 63.8%, 100% vs 54.6%, respectively, all p<0.001). They had greater problems with implementing infection control measures and were more likely to report feeling under pressure from managers or colleagues to be less cautious about infection control than they would like (60.0% vs 31.7%, p<0.001). Community staff reported greater difficulty in using new technologies in their work and in engaging service users with intellectual and other developmental disabilities in remote appointments.

#### Staff working only with people with intellectual disabilities and/or autism

Fifty eight respondents indicated that they worked only with people with intellectual disabilities or other neurodevelopmental disorders. The response patterns were similar to the whole sample in terms of items endorsed as very or extremely relevant.

## DISCUSSION

### Main results and implications

This is the first study to examine experiences of staff working across a range of mental healthcare settings for people with intellectual and other developmental disabilities in a pandemic. The survey data were collected at the peak of the pandemic, when lockdown restrictions were at their height and there was considerable uncertainty and anxiety around individual outcomes of infection and the implications of the pandemic for society as a whole. There is a notable lack of such information from previous pandemics; the only published paper specifically related to this group reported the implementation of infection control measures in an inpatient unit to reduce the risk of spreading SARS amongst those with intellectual disabilities (Wong et al. 2005).

Among staff concerns, the most salient were the implementation of safety measures to reduce the risk of transmission in in-patient care; the rapid adoption of remote working without adequate support and training compounded by difficulties in engaging service users in telehealth and remote appointments without prior experience (mainly for those in community settings); and gaps in care for this vulnerable population group due to the withdrawal or pause of usual sources of support. Although a significant proportion of staff reported resilience amongst service users and carers as a mitigating factor, we must remember that this survey was conducted relatively early in the first stage of the pandemic and that service users’ and carers’ material and psychological resources may be depleted as time progresses. The provision of comprehensive health and social care services for people with intellectual disability and other developmental disorders, following Government guidance and with mitigations to create a ‘COVID-secure’ environment, should be assured.

Half of survey respondents had significant concerns related to the challenges that people with cognitive impairments might have in understanding and responding to guidance around self isolation and social distancing, and having sufficient information in accessible formats. Similar difficulties may be shared by people with severe mental illness. Although a plethora of information has been developed using different media to explain the pandemic in accessible formats, the reach and effectiveness of these materials remains unclear.

Although clearly an unwelcome and unexpected situation, the COVID-19 pandemic prompted a number of rapid service changes that staff would like to see retained when the pandemic subsides, including practical changes and broad organisational and cultural shifts, such as a “*less authoritarian and more collaborative”* style from senior managers. Staff frequently remarked on the benefits of remote working whilst also recognising practical difficulties in its wide scale adoption, for both themselves and for service users. There are concerns that digital health interventions may not be suitably adapted for those with communication and cognitive impairments and may exclude them as a result, thus widening existing health inequalities (Sheehan and Hassiotis, 2017). Adequate information technology support for staff, maintaining dialogue between stakeholders, and following a co-production approach to the development of digital innovations will be important to ensure efficient and equitable services.

Staff concerns differed according to the sector and the setting within which they worked and it appears that those with managerial responsibility felt under more pressure than those without a management role. Staff sought support during the pandemic from a range of sources and turned to their employing organisation, managers, and peers for this, rating local support ahead of more generic national advice or wider initiatives. These findings indicate the need to tailor staff support according to local needs and highlight the importance of supporting staff at all levels of seniority.

Support from the public was ranked moderately highly amongst the whole sample and was rated as significantly more important by NHS staff compared with non-NHS workers. This corresponds with recent critique of high profile displays of appreciation for keyworkers, such as the weekly ‘Clap For Carers’, as being heavily skewed towards NHS staff, with those working in other sectors receiving relatively less recognition.

### Limitations

This is a secondary analysis of data from a larger survey and shares the limitations of the original study. It presents the perspective of staff who work with service users and, as such, is only one viewpoint, albeit direct, of what have been unprecedented changes. The sample is one of convenience; although the survey was open and respondents were drawn from a number of different sources it may not be representative of all staff working in mental healthcare for people with intellectual and other developmental disabilities. Respondents working in the NHS are over-represented compared with those working in other sectors. The questionnaire, whilst broad and devised by a multi-disciplinary team including experts by experience, was necessarily rapidly developed and it is possible that some important aspects of the pandemic and its impact on mental health services were not covered. Many other professionals important to the mental health care of people with intellectual disabilities and developmental disorders, such as those working in primary care and pharmacists, were not included in the sample. Future work should also aim to increase participation rates of non-white staff groups given what is now known about the impact of COVID-19 on their overall health and their importance as key workers (Patel et al. 2020).

### Conclusions

The COVID-19 pandemic has placed great strain on staff providing mental health services. The implications of the pandemic on mental health of staff and the population at large may not yet be fully apparent, with a ‘tsunami’ of mental health problems predicted over the coming months (Royal College of Psychiatrists, 2020). People with intellectual and other developmental disabilities face a unique set of challenges and may be especially affected. The impact on mental health services for this often marginalised group has remained under-reported.

It is vital to be prepared for future outbreaks of COVID-19 infections; in addition to ensuring the application of infection control methods (e.g. ensuring access to personal protective equipment) our survey results highlight the importance of developing contingencies that will allow for the continuation of services under possible future lockdown conditions. This includes establishing remote working solutions and effective telehealth interventions for people with intellectual and other developmental disabilities. An international collaboration investigating the specific impacts of COVID-19 across countries, and exploration of what works and what does not, are much needed in order to ensure that high-quality individualised care is available to vulnerable service users whose needs must not be an afterthought.

## Data Availability

The survey dataset is currently being used for additional research and is therefore not currently available in a data repository.

## Funding

This paper presents secondary analysis of independent research commissioned and funded by the National Institute for Health Research (NIHR) Policy Research Programme, conducted by the NIHR Policy Research Unit (PRU) in Mental Health. The views expressed are those of the authors and not necessarily those of the PRU, NIHR, the Department of Health and Social Care or its arm’s length bodies, or other government departments.

## Ethics approval

The King’s College London Research Ethics Committee approved this study (reference: MRA-19/20-18372).

## Consent to participate

Information on participation was provided on the front page of the survey. By starting the survey, participants agreed that they had read and understood all this information.

## Consent for publication

It was explained on the front page of the survey that responses may be used in articles published in scientific journals and that these articles will not include any information which could be used to identify any participant.

## Availability of data and material

The survey dataset is currently being used for additional research and is therefore not currently available in a data repository. A copy of the survey is available at this web address: https://opinio.ucl.ac.uk/s?s=67819.

## Conflict of interests

The authors declare no conflicts of interest.

## Notes

### Competing Interest Statement

The authors have declared no competing interest.

### Author Declarations

The Kings College London (UK) Research Ethics Committee approved this study (reference MRA-19/20-18372).

